# Age-specific control and Alzheimer’s disease reference curves and z-scores for glial fibrillary acidic protein in blood

**DOI:** 10.1101/2024.12.04.24318285

**Authors:** Steffen Halbgebauer, Badrieh Fazeli, Veronika Klose, Gabriele Nagel, Angela Rosenbohm, Dietrich Rothenbacher, Franziska Bachhuber, Sarah Jesse, Markus Otto, G. Bernhard Landwehrmeyer, Ahmed Abdelhak, Axel Petzold, Albert C. Ludolph, Hayrettin Tumani, the ALS Registry Swabia study group

## Abstract

**Background:** Serum glial fibrillary acidic protein (GFAP) is a biomarker for astrocytic injury and astrogliosis. Concentrations are elevated in numerous neurological disorders, including a pronounced increase in Alzheimer’s disease (AD). However, GFAP levels in the serum also increase with age. Consequently, the integration of GFAP levels into clinical routine and their interpretation demands age-adjusted reference values.

**Methods:** Serum from 1273 subjects (952 non-inflammatory and non-neurodegenerative neurological controls and 321 subjects with AD) were analyzed for GFAP using the microfluidic Ella system. Age-dependent serum GFAP reference values were calculated by additive quantile regression analysis. Percentiles and z-scores were employed for the presentation of GFAP levels as a function of age.

**Results:** All patients within the AD continuum exhibited statistically elevated serum GFAP levels in comparison to the control cohort (p<0.0001). This remained the case when the newly generated age-corrected z-scores were applied (p<0.0001). In the control cohort, a non-linear elevation of serum GFAP with increasing age was observed (Spearman correlation coefficient (r) 0.62, 95%CI 0.58-0.66, p<0.0001). In contrast, the AD cohort exhibited a more linear increase in serum GFAP levels (0.16, 95%CI 0.05-0.26, p=0.004). Age-dependent cut-offs for serum GFAP were calculated for different AD age groups. The calculated areas under the curve (AUC 0.97) demonstrated excellent diagnostic test performance in the early onset age group. This effect was less marked in the elderly subjects (AUC 0.72).

**Conclusions:** Our novel GFAP z-scores enable the integration and interpretation of serum GFAP levels in clinical practice, moving from group to individual level. They support both intra- and interindividual interpretation of single GFAP levels in neurological diseases with astrocytic pathology, including an accurate discrimination of Alzheimer’s disease.

## Introduction

Glial fibrillary acidic protein (GFAP) is a type III intermediate filament almost exclusively expressed in astrocytes in the central nervous system (CNS). GFAP is crucial for the mechanical strength of astrocytes and several of their functions such as the regulation of the blood-brain-barrier ^1^. In the context of astrogliosis, a process e.g. observed following neurodegeneration and neuronal death, there is an increase in GFAP expression. In addition to the normal turnover, following astrocytic injury, GFAP is released into the extracellular space, subsequently reaching the cerebrospinal fluid (CSF) and ultimately the bloodstream ^2^. In both matrices, CSF and blood, GFAP can be measured using different proteomics approaches, including mass spectrometry, as well as immunoassays such as ELISA, Simoa, and microfluidic assays like Ella ^3^. Given the expression pattern of GFAP, levels in the CSF are higher than in the blood, which presents a more challenging analytical environment due to strong matrix effects. Nevertheless, numerous studies have consistently demonstrated that blood GFAP exhibits superior discriminatory capabilities between diseases ^3–5^. It is hypothesized that this may be attributed to a partially direct release of GFAP through the astrocytic endfeet into blood vessels within the CNS ^6,7^. One neurological condition in which blood GFAP levels are markedly elevated is Alzheimer’s disease (AD) ^5,8,9^. Moreover, in genetic AD patients blood GFAP levels appear to increase by more than 10 years before the clinical onset of symptoms, suggesting that they may also have prognostic value ^10,11^. Additionally, studies have demonstrated its utility as a progression marker from AD with mild cognitive impairment (AD-MCI) to AD dementia ^12,13^. In therapeutic studies targeting Aβ, GFAP blood levels have been observed to decrease after several months of treatment, potentially reflecting a reduction in astrocytic damage or astrogliosis ^14,15^. Consequently, GFAP may also prove to be a highly valuable treatment monitoring marker in a clinical setting on an individual level. However, studies have consistently demonstrated that age is correlated with GFAP levels in both CSF and blood ^13,16–18^. This renders the interpretation of GFAP levels in clinical routine more challenging in the absence of age-dependent reference values.

In this study, nearly 1,000 serum samples of control patients without neuroinflammatory and neurodegenerative diseases from a broad age range were used to establish age-dependent reference curves, absolute values, and z-scores for serum GFAP. Additionally, the same methodology was applied to a cohort of over 300 AD samples, enabling the calculation of age-dependent cut-offs for the diagnosis of AD.

## Methods

### Patients

For this study, there were two sources for control patients. 1) The populations based ALS Swabian registry which also includes controls and 2) patients seen at the Department of Neurology at the University Hospital Ulm which were classified as controls (see below). From the population-based ALS Swabian register we analyzed 577 participants enrolled as controls which were sampled randomly from the general population (ethics votes No. 11/10, No. B-F-2010-062 and No. 7/11300). The study design and recruitment procedures of the ALS Swabian register have been described previously ^19–22^.

To make the additive quantile regression analysis for the generation of age-specific GFAP percentiles and z-scores more accurate we additionally measured samples from 424 patients seen at Ulm University Hospital between 2014 and 2023 (for selection and inclusion/exclusion criteria see flow chart Fig. S1). The patients were selected through convenience sampling. For the 424 patients seen at Ulm University Hospital acute neuroinflammation of the CNS was ruled out by CSF analysis (normal cell count, no evidence of intrathecal immunoglobulin synthesis). In addition, the patients did not show clinical or radiological signs for chronic neuroinflammation and neurodegeneration. For more details on the diagnoses refer to table S1 in the supplements. Due to GFAP levels below the lower limit of detection (LOD) 49 control subjects were excluded from further analysis.

The 324 AD patients were clinically diagnosed at Ulm University Hospital according to the IWG-2 criteria ^23^ and sampled between 2009 and 2023 (see flow chart Fig. S1). Additionally, all CSF AD samples were retrospectively examined for the ATN core markers (A: CSF Aβ1-42 to Aβ1-40 ratio, T: CSF phospho-tau 181 (p-tau181) and N: CSF total-tau (t-tau)), to be able to classify them according to the ATN system ^24^. All ADs were A+ with 270 patients A+T+ (270 A+T+N+ and 0 A+T+N-) and 51 A+T- (21 A+T-N+ and 30 A+T-N-) (three patients were excluded due to GFAP levels below the LOD). Control and AD patients with an acute or chronic renal insufficiency were excluded from the study.

The examination was approved by the local ethics committee (approval number Ulm 20/10) and conducted following the Declaration of Helsinki. All participants gave their written informed consent to participate in the study.

### Sampling and biomarker measurements

Blood samples were collected by venous sampling, centrifuged at 2000 g for 10min and the extracted serum aliquoted and frozen on the same day at −80° C. All serum samples were stored in polypropylene tubes.

For serum GFAP quantification we applied the microfluidic Ella platform (BioTechne, Minneapolis, USA) using the 2^nd^ generation GFAP cartridges, which were recently technically and clinically validated ^25^. The analyses were performed according to the manufacturer’s instructions. Intra- and interassay variations for two serum QCs measured in duplicates on each cartridge, were below 20%.

ATN markers were analyzed using the Lumipulse G 600II platform (Fujirebio, Tokyo, Japan).

### Statistics

The distribution of data was assessed visually and statistically. Because data were non-Gaussian data, non-parametric tests were used.

Due to a nonlinear relation of age and serum GFAP levels additive quantile regression analysis was performed based on the control population to assess the effect of age on GFAP concentrations. According to this analysis we determined the z-scores for the control group. Furthermore, we also applied this model to calculate the AD z-scores.

For a two-group comparison the Mann-Whitney-U Test (two-tailed), for more groups the Kruskal-Wallis test followed by Dunn’s multiple comparisons test was applied with a p<0.05 indicative of statistically significant results. For the discrimination controls vs. AD and the calculation of cut-offs receiver operating characteristic (ROC) analysis was applied. The Youden’s Index was calculated for optimization of the cut-off levels. For association testing between serum GFAP and other parameters Spearman rank correlations were applied. The visualization and analysis was performed with RStudio V. 4.3.1 and GraphPad Prism V.10.3.1 (GraphPad, Software, La Jolla, California, USA).

## Results

The demographic as well as GFAP serum values of the control and AD cohort are shown in Table 1. For the establishment of age-dependent control and AD GFAP reference curves and values 952 controls and 321 AD patient samples were used.

**Table 1.**
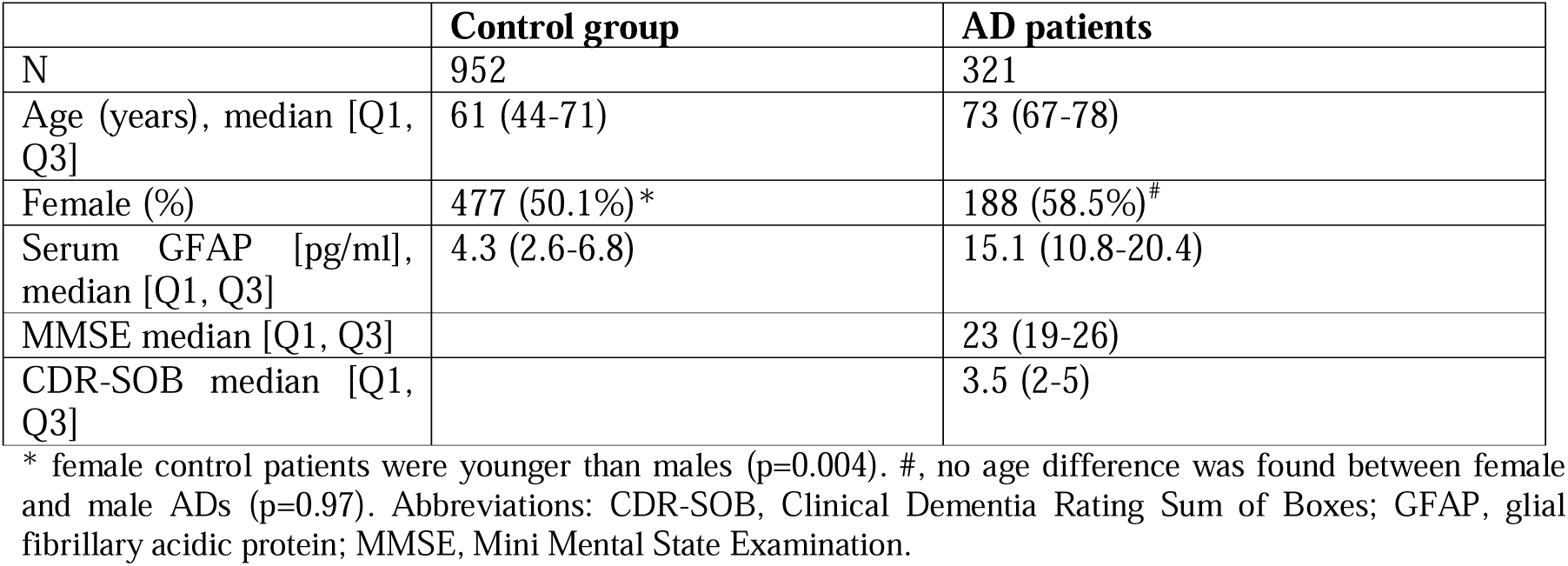
Study cohort. Data are shown as median, interquartile range, N and percentage (%).

### Serum GFAP levels in the control cohort

Figure 1A depicts the GFAP serum reference curves from the 25^th^ to the 95^th^ percentile in the control cohort, stratified by age. The serum GFAP values demonstrate an increase with age, commencing at approximately 50 years. The median (interquartile range) for the group of patients below 50 years was 2.6 pg/ml (1.8-3.6 pg/ml), while the patients above 80 years of age exhibited a significantly higher median of 11.7 pg/ml (7.1-17.0 pg/ml). The correlation between age and serum GFAP levels was found to be moderate to strong, with a Spearman r of 0.62 (95% CI 0.58-0.66), p<0.0001. Table 2 presents the serum GFAP concentrations corresponding to the 50^th^ and 95^th^ percentiles for various age groups. Figure 1B presents the z-score age reference curves rather than percentiles. Z-score values can be found in table S2. We found no significant difference between serum GFAP levels in female and male control patients when looking at the whole control group (p=0.07). Results for GFAP levels stratified by age and sex can be found in the supplementary materials.

**Figure 1:**
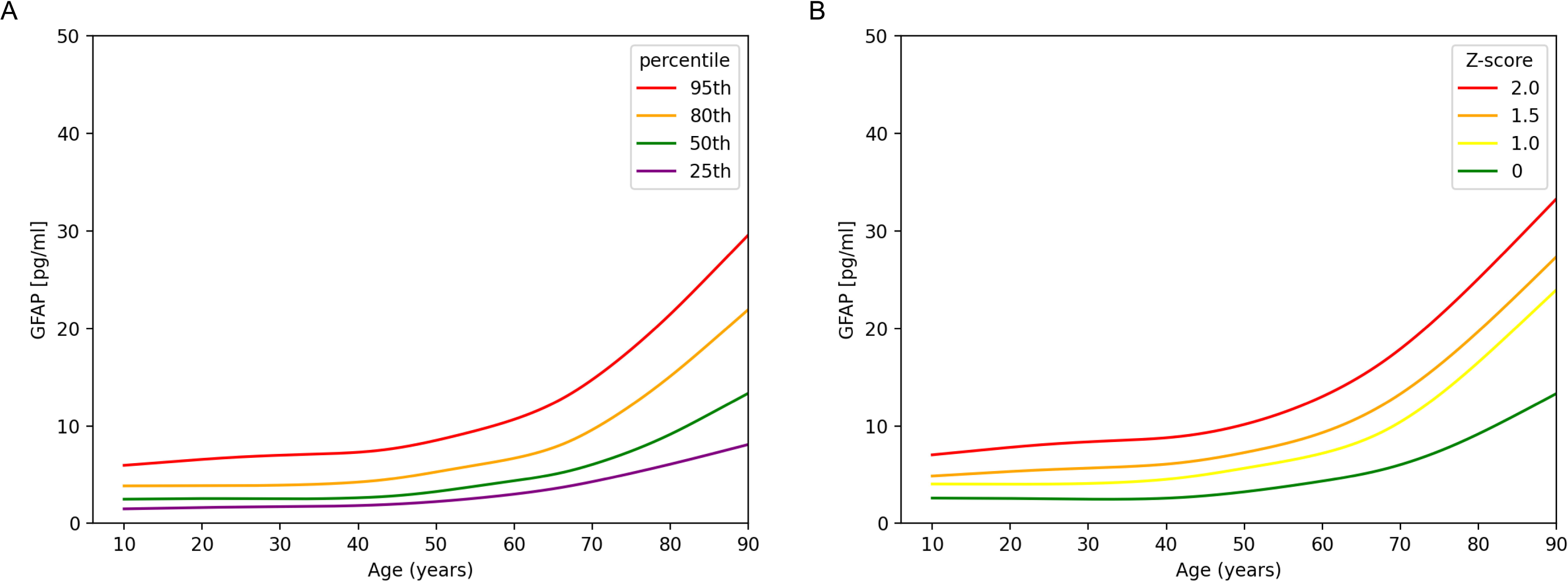
Serum GFAP age-dependent control reference curves. **A** displays the serum GFAP percentiles dependent on age. In **B** the z-scores from z=0 (corresponding to the 50^th^ percentile) until z=2 are illustrated. For the modelling additive quantile regression analysis of 951 control patients was applied. Abbreviations: GFAP; glial fibrillary acidic protein

**Table 2:** Age-specific 50% and 95% GFAP percentiles in serum of control and AD patients:

| Age (Years) | Control 50th percentile [pg/ml] | Control 95th percentile [pg/ml] | AD 50th percentile [pg/ml] | AD 95th percentile [pg/ml] |
| --- | --- | --- | --- | --- |
| 20 | 2.48 | 6.07 | n/a | n/a |
| 25 | 2.49 | 6.37 | n/a | n/a |
| 30 | 2.51 | 6.67 | n/a | n/a |
| 35 | 2.55 | 6.97 | n/a | n/a |
| 40 | 2.66 | 7.32 | 10.8 | 27.6 |
| 45 | 2.87 | 7.75 | 11.4 | 28.3 |
| 50 | 3.24 | 8.38 | 12.1 | 29.0 |
| 55 | 3.77 | 9.27 | 12.8 | 29.6 |
| 60 | 4.35 | 10.4 | 13.5 | 30.3 |
| 65 | 5.00 | 12.0 | 14.1 | 30.9 |
| 70 | 6.03 | 14.4 | 14.8 | 31.6 |
| 75 | 7.32 | 17.6 | 15.5 | 32.2 |
| 80 | 8.95 | 21.4 | 16.1 | 32.9 |
| 85 | 11.4 | 26.5 | 16.8 | 33.5 |

Abbreviations: AD, Alzheimer's disease; GFAP, Glial fibrillary acidic protein

### Serum GFAP levels in the AD cohort

In the AD cohort GFAP values were significantly increased compared to controls in all AD, ATN+ ADs and ADs with a A+T- (N+/-) CSF biomarker profile (Figure 2A and B). We detected no difference between the AD groups. Figure 2C illustrates that GFAP values were also elevated in the different AD groups when age corrected z-scores are applied. For the discrimination between control and AD patients ROC analysis depicted an AUC of 0.93 (95%CI 0.91-0.94) for all ADs, 0.93 (95%CI 0.92-0.95) for the AD A+T+N+ and 0.92 (95%CI 0.89-0.95) for the AD A+T-(N+/-) group (Figure 2D). The optimal cut-off for the whole AD group was determined to be at 8.1 pg/ml with a diagnostic sensitivity of 92% (95%CI: 88-94%) and specificity of 84% (95%CI: 80-85%).

**Figure 2:**
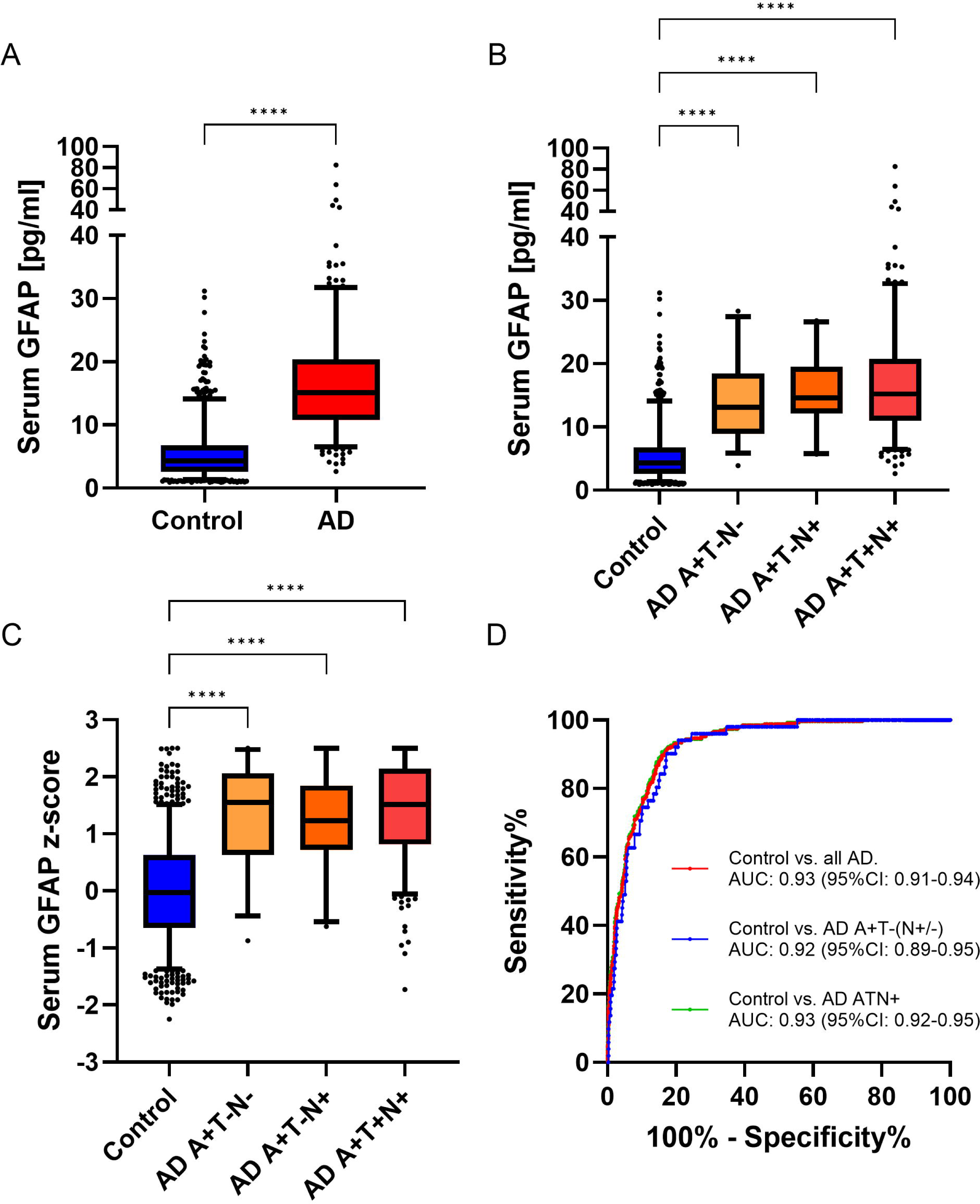
Serum GFAP analysis in the AD cohort. **(A)** shows the serum GFAP comparison between control and all AD patients with significant higher levels in the AD group. (**B**) depicts the comparisons with the AD group stratified according to CSF ATN markers. All groups demonstrated markedly increased serum GFAP concentrations compared to controls. The same is true when age-corrected z-scores (**C**) are applied. In **D** the discriminating potential of serum GFAP for controls vs all ADs, AD A+T+ and A+T- is illustrated. The ROC analysis yielded nearly the same high AUCs for all comparisons. Abbreviations: AD; Alzheimer’s disease; GFAP; glial fibrillary acidic protein; ROC, Receiver operating characteristics

Moreover, the availability of control samples across a wide age range enabled a comparison of serum GFAP concentrations across different age groups (Figure 3A). All AD patient groups between the ages of 50 and 90 exhibited significantly elevated levels in comparison to the corresponding age control group. The results of the ROC analysis indicated that the youngest patient group with ADs and controls between the ages of 51 and 60 exhibited the highest AUCs. Subsequently, the area under the curve (AUC) values decreased with increasing age of the stratified age groups (Figure 3B). The optimal cut-off value for each age group, along with the corresponding sensitivity and specificity values, can be found in the supplementary materials (Table S4).

**Figure 3:**
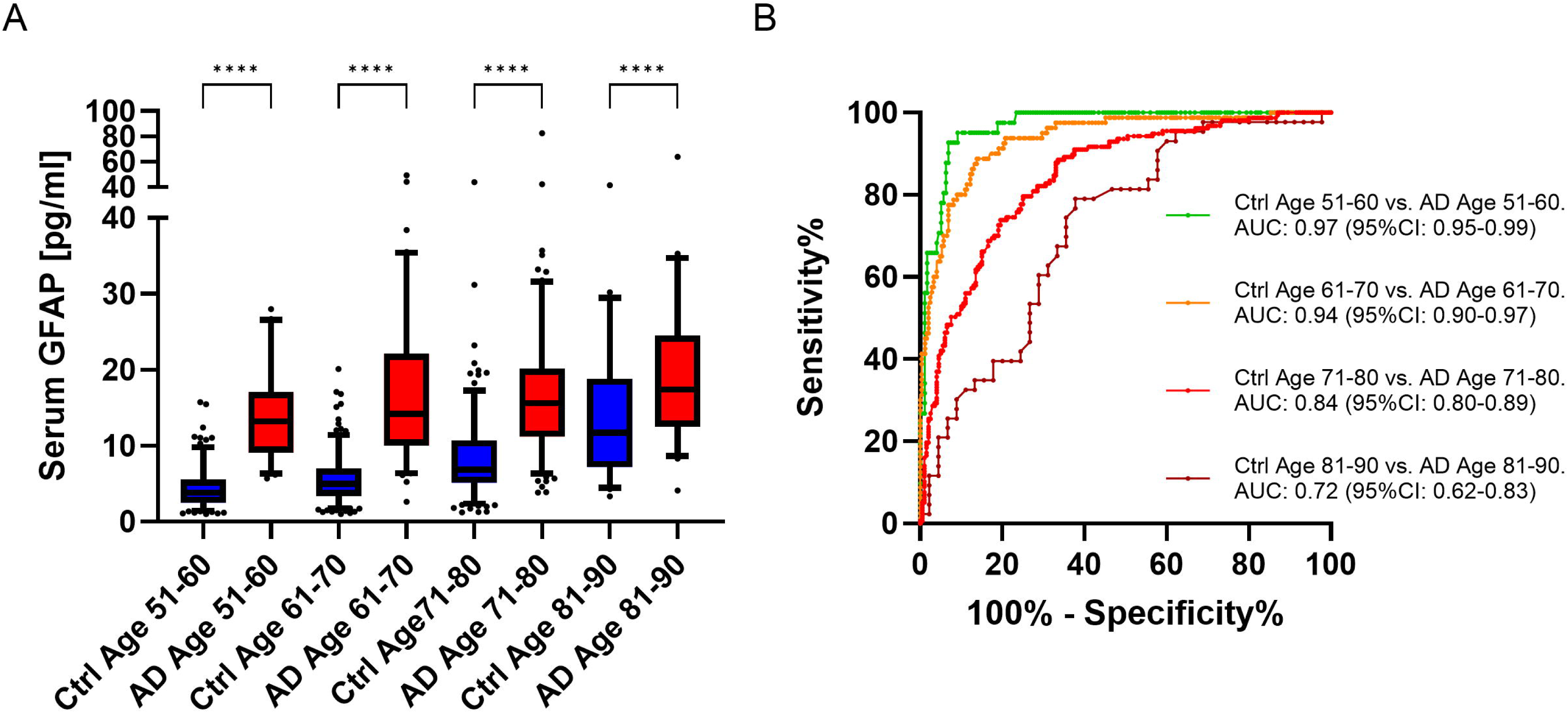
Comparison of serum GFAP stratified by age groups of 10 years. In (**A**) serum GFAP levels stratified according to age decade are compared between control and AD patients. All age groups show significantly increased levels in AD. (B) depicts the serum GFAP ROC analysis between the control and corresponding AD age groups. The AUCs are highest in the younger age groups and decline with older age. Abbreviations: AD; Alzheimer’s disease; Ctrl, control; GFAP; glial fibrillary acidic protein; ROC, Receiver operator characteristics

Subsequently, we conducted a more detailed examination of how age affects GFAP levels within the AD cohort. Figure 4A illustrates the linear elevation of GFAP concentrations with increasing age, as demonstrated by quantile regression analysis. Table 2 presents the serum GFAP concentrations for the 50th and 95th percentiles for different age groups in the AD cohort. The correlation coefficient for the GFAP and age in the AD cohort was determined to be 0.16 (95%CI 0.05-0.26), p=0.004. Figure 4B and table S2 display the corresponding z-scores. In the AD group there was a trend to higher levels in female patients which was, however, not statistically significant in the whole cohort (p=0.07) and when stratified according to age (see supplementary materials).

**Figure 4:**
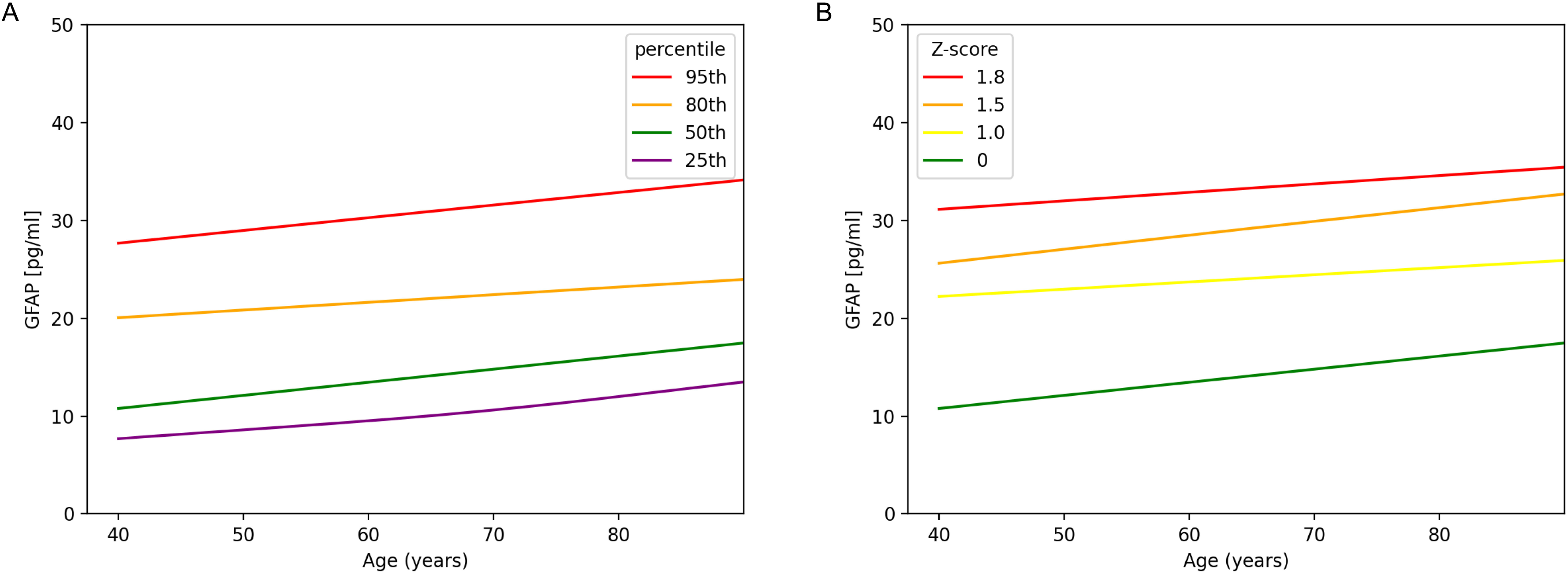
Serum GFAP age-dependent AD reference curves. **A** displays the serum GFAP percentiles dependent on age of the AD patient cohort. In **B** the z-scores from z=0 (corresponding to the 50^th^ percentile) until z=1.8 are illustrated. For the modelling additive quantile regression analysis was applied. Abbreviations: GFAP; glial fibrillary acidic protein

### Correlations with ATN markers and cognitive scores

The correlation between serum GFAP levels and CSF t-tau (0.14, 95%CI 0.03-0.25, p=0.011) and p-tau 181 (0.14, 95%CI 0.03-0.24, p=0.012) in the AD group was found to be weak. A moderately stronger correlation was identified with the cognitive scores MMSE (−0.27, 95%CI −0.39--0.14, p<0.0001) and CDR-SOB (0.22, 95%CI 0.03-0.40, p=0.01) (see also Figure S5 in the supplementary materials). No significant differences were observed in CSF t-tau and p-tau levels across the various age groups (Figure S6 A-B). In contrast, MMSE scores and CDR-SOB values exhibited a decline or increase, respectively, in the oldest age group (see supplementary figure S6 C-D).

When the AD cohort was divided into patients with MCI (AD-MCI) and dementia (ADD) serum GFAP levels were already elevated in the MCI group compared to controls (p<0.0001) and remained elevated in the ADD group (p<0.0001) (see supplementary materials figure S7).

## Discussion

Serum GFAP is an important fluid astrocytic biomarker that is increasingly being recognized as a valuable tool for routine applications in clinical settings. Nevertheless, in order to interpret serum GFAP levels in routine analysis, it is of utmost importance to account for the observed increase in levels with advancing age, a phenomenon that has been consistently demonstrated in numerous publications. This study addresses this issue by calculating and graphically displaying age-corrected reference values using absolute values and z-scores. Given the elevated serum GFAP levels observed in AD patients in the literature, we also determined age-reference values for AD and calculated age-dependent cut-off levels.

The results demonstrate a clear increase in serum GFAP levels with age, which is nevertheless less pronounced than that observed for the neurofilament light chain protein (NfL), for which several age-reference studies have been conducted ^26–28^. The available data on serum GFAP reference values, however, is limited. Studies with a smaller number of adults conducted by Danish and Canadian colleagues examined GFAP reference values using the Simoa technology ^29,30^. They also demonstrate an increase with age starting around 50 years of age, which is less pronounced than for NfL. It should be noted, however, that the absolute values of these studies and our data are not directly comparable, and no z-scores were reported. The application of z-scores, calculated in our study, facilitates the interpretation of the data and renders it independent of the platform utilized to measure serum GFAP levels. Additionally, the use of age-corrected z-scores for GFAP, defined as the number of standard deviations a single GFAP value is above or below the mean GFAP level for a given age, offers further advantages, including a normal distribution and the potential for negative values.

The significance of age-reference values for the interpretation of GFAP levels can be illustrated by a straightforward example. According to our data, a serum GFAP measurement of 6 pg/ml is considered normal for patients at age 70 and elevated for patients at 20 years of age. The use of z-scores allows for the direct observation of this distinction without the need for a table or graphic. For instance, a serum GFAP value of 6 pg/ml corresponds to a z-score of −0.02 for an age of 70, indicating that the serum GFAP value of 6 pg/ml is nearly the mean of this age stage. However, a serum GFAP concentration of 6 pg/ml corresponds to a z-score of 1.63 at the age of 20, indicating a level that is more than one and a half times the standard deviation above the serum GFAP mean level at this age rendering it clearly elevated.

The GFAP analysis in the AD cohort corroborates the findings of previous studies which have demonstrated that serum GFAP levels are significantly elevated in AD patients compared to controls^5,8,9^. In addition to elevated levels in ATN positive AD patients, we found a significant increase also in AD patients only positive for the CSF beta amyloid 42 to 40 ratio. This finding is consistent with the results of previous studies that also identified elevated serum GFAP levels in A+T-patients ^4,16^. Furthermore, we demonstrate that the elevation in serum GFAP observed in AD patients is confirmed when age-corrected z-scores are applied. The observed AUCs between 0.72 and 0.97, depending on the age group, confirm the literature, which reports AUCs between 0.79 and 0.93 ^8,13,31^. The analysis of a large number of control patients across a wide age range enabled the establishment of age-specific cut-offs for AD. They can assist in differentiating between elevated serum GFAP levels resulting from disease-specific astrocytic injury or astrocytosis in AD and those caused by normal ageing effects in the elderly population.

Our findings for the age-reference curves for the AD cohort indicated a more linear increase compared to the control curves. In the literature there a no serum GFAP reference curves available for AD to compare with. However, NfL was also examined in AD, and a similar pattern of a more gradual linear increase was observed ^28^. In a disease cohort such as AD, it is necessary to determine whether the elevation of serum GFAP with increasing age is due to the effects of aging or to a more severe disease pathology in older age groups, which may result in increased GFAP levels in the blood. To address this question, we analyzed the correlation between serum GFAP and CSF markers, particularly total and phosphorylated tau, which are known to be associated with atrophy and disease intensity ^32–34^. Additionally, we evaluated the correlation between serum GFAP and cognitive scores, which provide insight into the degree of cognitive impairment. The weak correlation between serum GFAP and CSF t-tau and p-tau is in accordance with the findings of other studies, which indicate that serum GFAP is not a marker of tau pathology ^35^, In any case, the CSF t-tau and p-tau181 levels are not different between younger and older AD patients, indicating a degenerative process of comparable severity in the different age stages. However, the observed correlation with cognitive scores, which has also been documented in several other studies ^9,12,13,36^ indicates that serum GFAP may be associated with the severity of cognitive dysfunction in our AD cohort. The cognitive decline analyzed by MMSE and CDR-SOB also appears to be more pronounced in the oldest AD patients. Together, synaptic loss, the strongest pathological correlate of cognitive decline ^37,38^ and the effects of aging may contribute to the elevated GFAP levels observed in older AD patients.

The principal strength of our study is the analysis of together nearly 1,000 control subjects for the establishment of age-dependent reference values and z-scores. Furthermore, the generation of z-scores facilitates straightforward interpretation of the results and renders the interpretation independent of the analytical platform. Furthermore, the ATN-characterized AD group permitted the generation of age-specific cut-offs, which could prove invaluable in clinical routine analysis. A potential limitation of this study is the inapplicability of the results for patients with renal dysfunction and the relatively small subgroups of AD patients included. Future studies could aim to recruit a larger number of AD patients to improve the accuracy of very high or low z-scores.

In conclusion, our study offers age-dependent reference curves, values and z-scores for serum GFAP, which could greatly aid in clinical practice by supporting the interpretation of individual GFAP levels and facilitating the integration of GFAP analysis into clinical report. The reference values are applicable to any clinical scenario exploring active astrocytic changes in neurological diseases. Additionally, we provide age-specific serum GFAP cut-offs tailored for ATN-categorized AD patients.

## Acknowledgments

We thank the Ilona Kraft-Overbeck, Ines Dobias and Nicola Lämmle for their excellent field work, and Gertrud Feike, Sarah Enderle and Birgit Och for their excellent data management and technical support as well as all patients and healthy controls for participation in the study.

## Author Contributions

All authors made substantial contributions to conception and design, and/or acquisition of data, and/or analysis and interpretation of data.

All authors gave final approval of the version to be submitted and agree to be accountable for all aspects of the work in ensuring that questions related to the accuracy or integrity of any part of the work are appropriately investigated and resolved.

Conception and design of the study: SH, HT; Sample collection and data management: SH, BF, VK, GN, AR, DR, FB, SJ, MO, GBL, AA, AP, ACL, HT. Study management and coordination: SH, HT; Statistical methods and analysis: SH, BF, VK, HT; Interpretation of results: SH, BF, MO, GBL, ACL, HT; Manuscript writing (first draft): SH, HT; Critical revision of the manuscript: SH, BF, VK, GN, AR, DR, FB, SJ, MO, GBL, AA, AP, ACL, HT.

## Data availability

The data that support the findings of this study are available from the corresponding author upon reasonable request.

