## Supplementary materials for "Age-specific control and Alzheimer’s disease reference curves and z-scores for glial fibrillary acidic protein in blood"

### Supplementary tables and figures

Table S1: Diagnoses of the Ulm University Hospital control group

| Diagnosis | Number of patients |
|---|---|
| Acoustic hallucinations | 1 |
| Adjustment disorder | 1 |
| Allergic reaction | 1 |
| Analgesic-induced headache | 1 |
| anterior ischaemic optic neuropathy | 1 |
| Barbiturate intoxication | 1 |
| Bladder and rectal dysfunction (psychogenic) | 1 |
| Bladder atony of unknown cause | 1 |
| Capsulitis and tenditis of the right shoulder | 1 |
| Cervical spine syndrome with cervical myalgia (excluded meningitis) | 1 |
| Cervical spine pain syndrome | 1 |
| Chronic left-sided lumbalgia | 1 |
| Chronic myalgic syndrome, unclear cause | 1 |
| Clinical gait uncertainty | 1 |
| Cluster headache | 1 |
| Degenerative cervical spine syndrome with disc protrusion | 1 |
| Deregulated insulin-dependent diabetes mellitus | 1 |
| Discrete ptosis on the left | 1 |
| Distal symmetric axonal sensorimotor polyneuropathie | 1 |
| Dizziness | 1 |
| Exclusion diagnosis of ocular myasthenia | 1 |
| Exclusion of an organic cause for left-sided facial discomfort | 1 |
| Exclusion of inflammatory genesis of diffuse tingling paraesthesia | 1 |
| Exclusion of trigeminal nerve affection | 1 |
| Exercise-induced myalgias (muscle biopsy no myopathy) | 1 |
| Head and facial pain | 1 |
| Hemicrania continua with left-sided headache and Horner's syndrome left | 1 |
| Hemiplegic migraine | 1 |
| hip contusion | 1 |
| Hyp- and paraesthesia upper and lower extremities | 1 |
| hypaesthesia left | 1 |
| Hypaesthesia of the right arm of unclear aetiology | 1 |
| Hypaesthesia of the right leg of dissociative origin | 1 |
| Hypaesthesia of the right side of the neck and right upper arm | 1 |
| Hypaesthesia V2 and V3 left side of the face, exclusion of chronic inflammation | 1 |
| Hypothyroidism | 1 |
| Idiopathic abducens palsy on the right | 1 |
| Idiopathic sudden deafness on the right | 1 |
| Intermittent brachial and facial hypaesthesia without evidence of an organic cause | 1 |
| Left frontal headache, excluding secondary cause of headache | 1 |
| left-sided central sensorimotor hemisymptomatics | 1 |
| Lumbago | 1 |
| Medication-induced headache, dizzines | 1 |
| Migraine with temporary hemiparesis | 1 |
| Monarthritis right knee | 1 |
| Mononeuritis multiplex | 1 |
| Motor arm plexus palsy | 1 |
| Multifactorial gait disorder | 1 |
| Neuralgiform headache | 1 |
| No evidence of meningitis | 1 |
| No evidence of neuroborreliosis, erythema migrans after tick bite | 1 |
| Non-specific multilocular tingling paraesthesia | 1 |
| Non-specific paraesthesia of all extremities | 1 |
| Non-specific sensory disturbance of the right arm | 1 |
| Occipital neuralgia | 1 |
| Opressive headaches | 1 |
| OSG supination trauma | 1 |
| Pain in right hand of unknown origin | 1 |
| Pansinusitis | 1 |
| Parainfectious headache, Covid infection | 1 |
| Parese des N. peronaeus profundu | 1 |

| | |
|---|---|
| Physiological anisocoria (right < left), tension headache (exclusion ischemia, tumor) | 1 |
| Pressure lesion of the lower brachial plexus on the left | 1 |
| Presyncope | 1 |
| Primary somatoform dizziness | 1 |
| Prolong migraine with aura | 1 |
| Proximally emphasised arm paresis | 1 |
| Recurrent left-sided brachial and facial paresthesias | 1 |
| Recurrent orthostatic syncope | 1 |
| Recurrent polyradiculopathies of the left upper extremity | 1 |
| Regredient vertigo and tingling paraesthesia of both legs | 1 |
| Residual complaints metatarsophalangeal joint on the right | 1 |
| rheumatoid arthritis | 1 |
| Right abducens nerve palsy | 1 |
| Right trigeminal nerve affection | 1 |
| Right-sided tingling paraesthesia | 1 |
| S1 syndrome left | 1 |
| Sensible C6-syndrom | 1 |
| Sinusitis | 1 |
| Stress reaction | 1 |
| Subjective gait disturbance, no evidence of organic genesis | 1 |
| Subjective left sensorimotor hemisymptomatics, exclusion of central genesis | 1 |
| Suspected presyncope | 1 |
| Symptomatic facial pain on the left of dentogenic origin | 1 |
| Temporal arteritis | 1 |
| Tic disorder of unclear aetiology | 1 |
| Tingling paraesthesia | 1 |
| Tingling paraesthesia and neuropathic pain of the lower extremities | 1 |
| Tingling paraesthesia and pain in the left side of the body | 1 |
| Tingling paraesthesia of unclear aetiology | 1 |
| Tingling Paresthesias in the extremitis without organic correlate | 1 |
| Transient Double images | 1 |
| Transient Hemihypaesthesia | 1 |
| Transient monoparesis of the left arm | 1 |
| Transient Tingling parasthesias | 1 |
| Transient vertigo | 1 |
| Unsystematic feelings of tingling and numbness | 1 |
| Unsystematic sensory disturbance on the left elbow and the soles of the feet | 1 |
| Unsystematic sensory disturbances of unclear cause | 1 |
| Vasovagal syncope in postural tachycardia syndrome | 1 |
| Ventricular extrasystole | 1 |
| Anxiety disorder | 2 |
| Benign fasciculations | 2 |
| brachial plexus neuritis | 2 |
| Choreatic hyperkinesia | 2 |
| Exclusion of a neuromuscular disease | 2 |
| Lumboischalgia | 2 |
| Meniere's disease | 2 |
| Migraine (atypical) | 2 |
| Migraine with vestibular aura | 2 |
| Nerve root affection | 2 |
| Oculomotor nerve palsy | 2 |
| Pain disorder | 2 |
| Papilloedema | 2 |
| Paraesthesia of the hands and feet on both sides | 2 |
| Post-puncture headache, tension headache | 2 |
| Pseudoradicular cervical syndrome | 2 |
| Tingling paraesthesia of the right extremities | 2 |
| Trigeminal autonomous headache | 2 |
| Atypical facial pain | 3 |
| Diffuse hypaesthesia | 3 |
| Episodic tension headache | 3 |
| hemihypaesthesia | 3 |
| Myalgia | 3 |
| Orthostatic synkope | 3 |
| Phobic vertigo | 3 |
| Recurrent presyncope | 3 |
| Functional leg paraparesis | 4 |
| Hypertension | 4 |
| Transient Sensory disturbance | 4 |
| Unsystematic dizziness | 4 |

| | |
|---|---|
| benign paroxysmal positional vertigo | 5 |
| Ocular Myasthenia | 5 |
| Sensory disturbance | 5 |
| Trigeminal neuralgia | 5 |
| Trochlear nerve palsy | 5 |
| Chronic pain disorder | 7 |
| Transient global amnesia | 7 |
| Depressive disorder | 8 |
| somatoform disorder | 9 |
| Facial palsy | 10 |
| Migraine | 10 |
| Somatisation disorder | 11 |
| Dissociative disorder | 13 |
| Migraine with aura | 13 |
| Exclusion of a neuroinflammatory CNS disease | 23 |
| Vestibular neuritis | 19 |
| Tension headache | 59 |

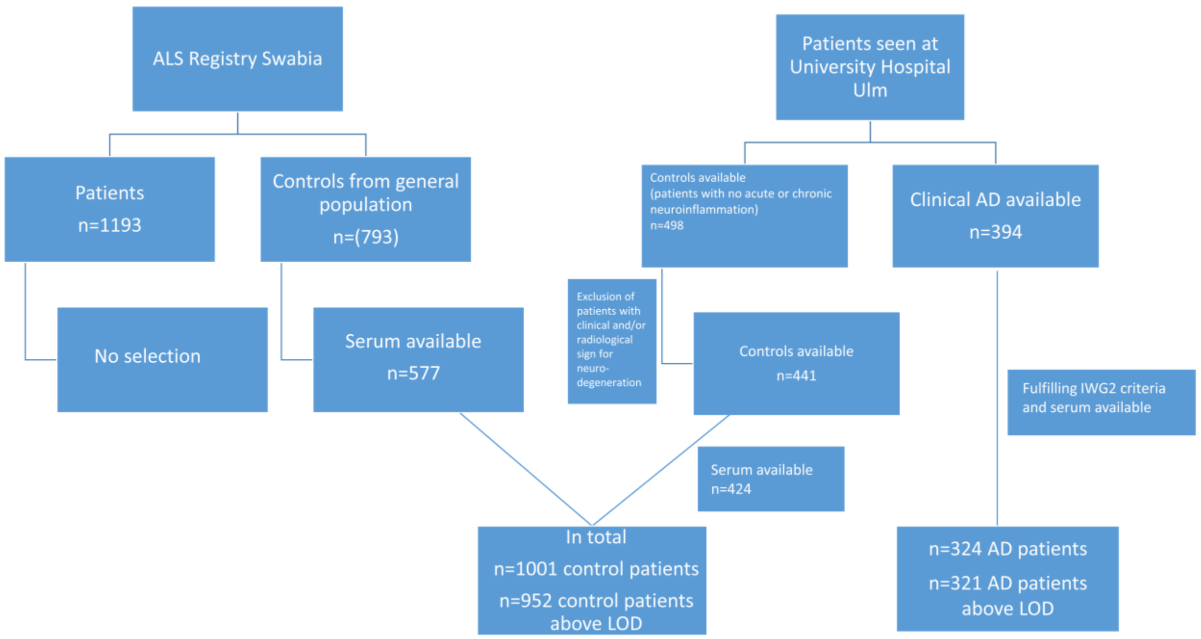


**Figure S1: Patient selection flow chart**

The figure displays the selection of control patients from the population-based ALS Registry Swabia and the selection of further controls and the AD cohort from patients seen at the University Hospital Ulm. From the 1001 measured control patients 49 had levels below the LOD and were excluded from further analysis. In addition, three ADs were excluded. Abbreviations: AD, Alzheimer's disease; n, number.

Table S2: Age-specific GFAP z-scores in serum of control and AD patients:

| Age | Control z=0 [pg/ml] | Control z=2 [pg/ml] | AD z=0 [pg/ml] | AD z=1.8 [pg/ml] |
|---|---|---|---|---|
| 20 | 2.48 | 8.15 | n/a | n/a |
| 25 | 2.49 | 8.51 | n/a | n/a |
| 30 | 2.51 | 8.88 | n/a | n/a |
| 35 | 2.55 | 9.24 | n/a | n/a |
| 40 | 2.66 | 9.63 | 10.8 | 31.2 |
| 45 | 2.87 | 10.1 | 11.4 | 31.6 |
| 50 | 3.24 | 10.8 | 12.1 | 32.0 |
| 55 | 3.77 | 11.8 | 12.8 | 32.4 |
| 60 | 4.35 | 13.1 | 13.5 | 32.9 |
| 65 | 5.00 | 15.0 | 14.1 | 33.3 |
| 70 | 6.03 | 17.8 | 14.8 | 33.7 |
| 75 | 7.32 | 21.4 | 15.5 | 34.2 |
| 80 | 8.95 | 25.4 | 16.1 | 34.6 |
| 85 | 11.4 | 29.9 | 16.8 | 35.0 |

Abbreviations: AD, Alzheimer's disease; Con, control; GFAP, Glial fibrillary acidic protein

**Stratification of serum GFAP levels according to sex and age**

In the age group of 50 we found significantly elevated serum GFAP levels in females compared to males in the control cohort (Fig. S2 A and B). This trend started with 40 until 89 years of age. In AD there was a trend to higher levels in female compared to male patients in the whole cohort (p=0.07). When we analyzed GFAP levels in the AD cohort according to age we didn't detect a statistically significant sex difference (Fig. S3 A and B).

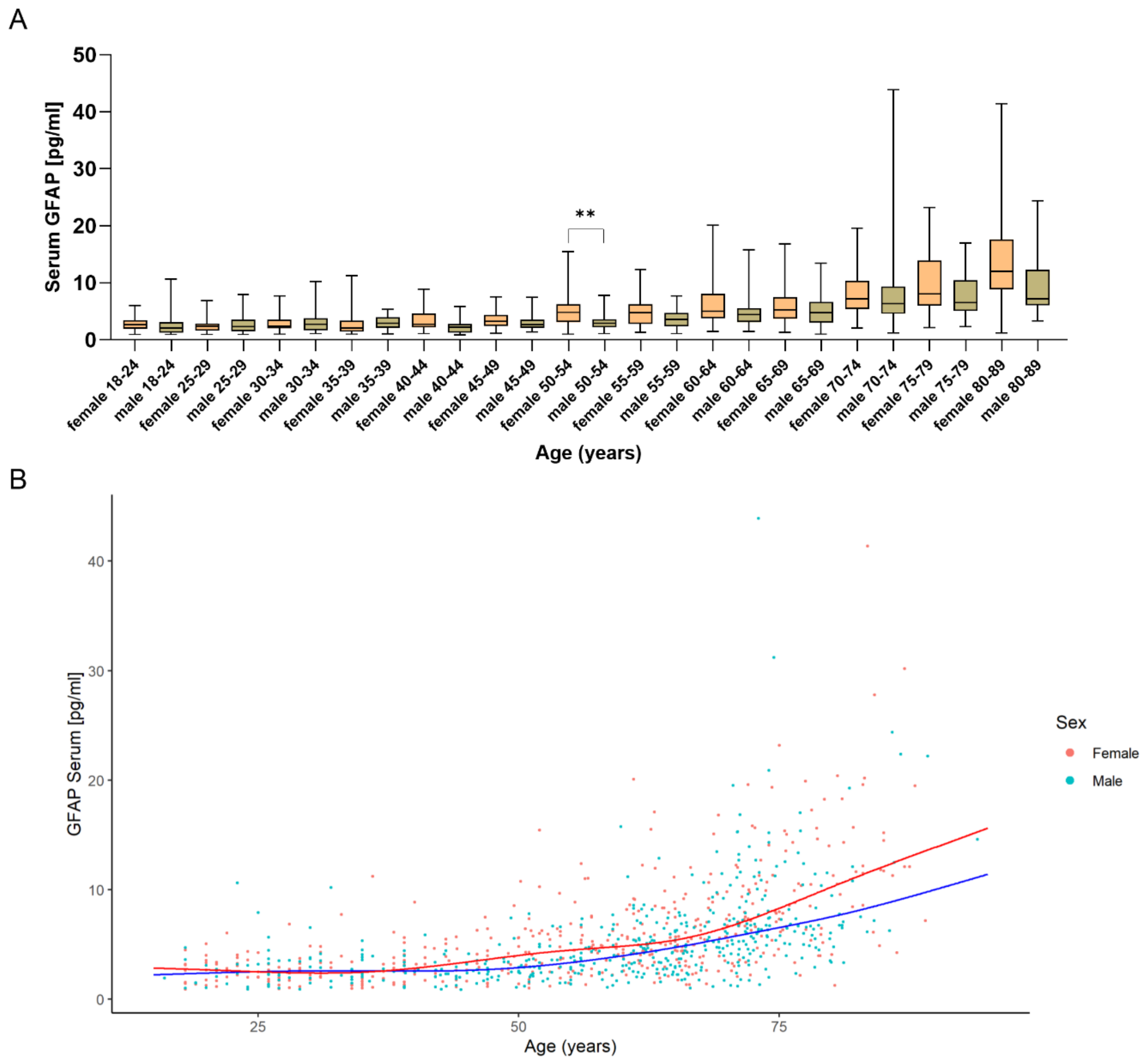


**Figure S2: GFAP levels according to age and sex in the control cohort**

Figure S2 **A** displays GFAP levels in female and male control patients according to age. A trend to higher levels starting with 40 years of age can be seen. In **B** the median of female (red) and male control subjects according to additive quantile regression is plotted. Differences between female and male GFAP levels in the corresponding age groups (planned comparisons) were compared using Kruskal-Wallis not corrected for multiple comparisons (Uncorrected Dunn's test). **, $p<0.01$. Abbreviations: GFAP, glial fibrillary acidic protein

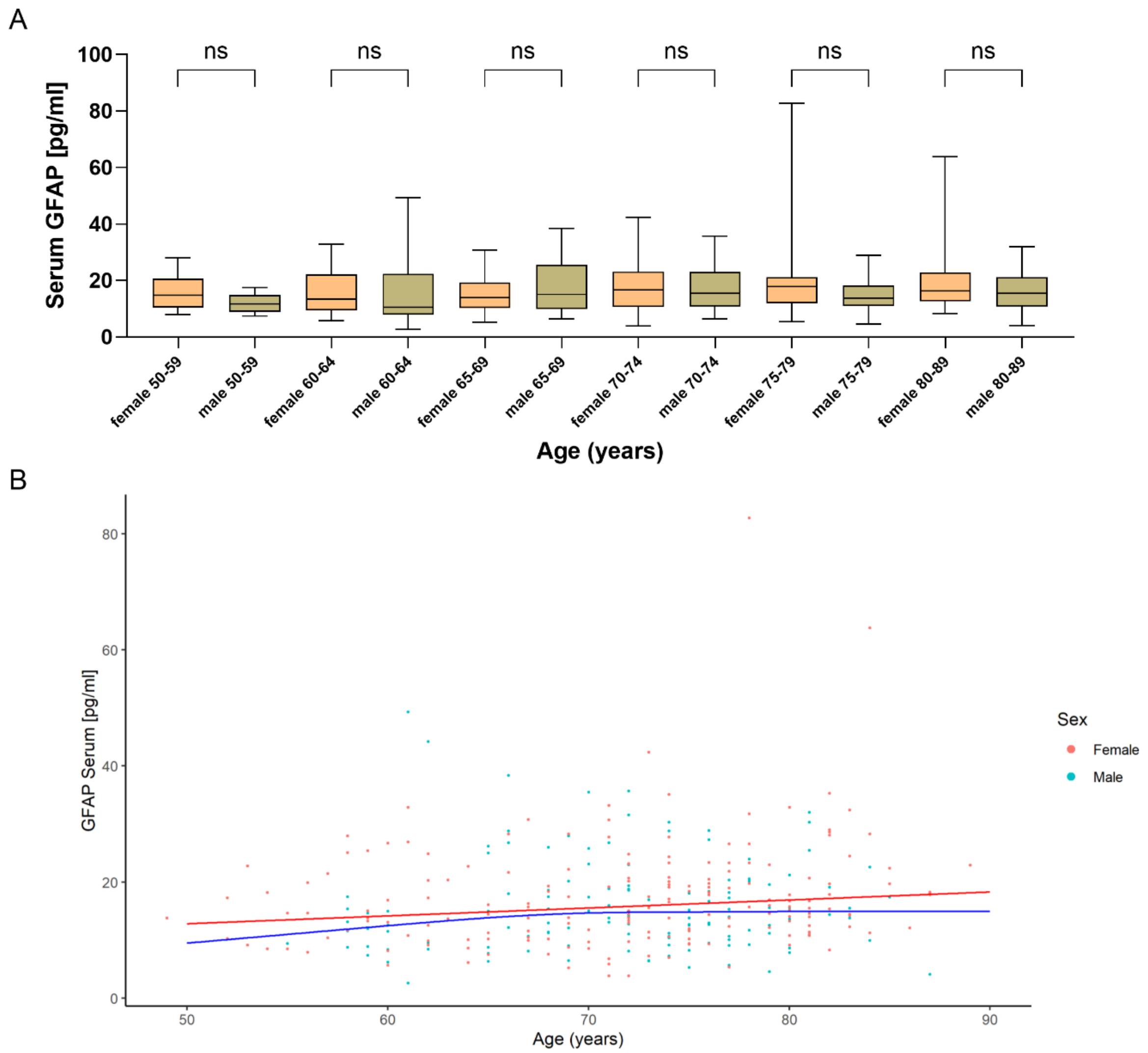


**Figure S3: GFAP levels according to age and sex in the AD cohort**

Figure S3 **A** displays GFAP levels in female and male control patients according to age. No significant difference was detected. In **B** the median of female (red) and male AD subjects according to additive quantile regression is plotted. Differences between female and male GFAP levels in the corresponding age groups (planned comparisons) were compared using Kruskal-Wallis not corrected for multiple comparisons (Uncorrected Dunn's test). Abbreviations: GFAP, glial fibrillary acidic protein

We therefore performed a separate additive regression analysis for female and males control subjects in the control cohort and plotted the percentiles (Fig. S4A-B) and z-scores (Fig. S4C-D) starting from 40 years of age. In table S3 the corresponding z-score values for z=0 and z=2 can be found. In mean the female z-score 0 and 2 values from 40 years onwards are 20 and 12 % higher than in males, respectively.

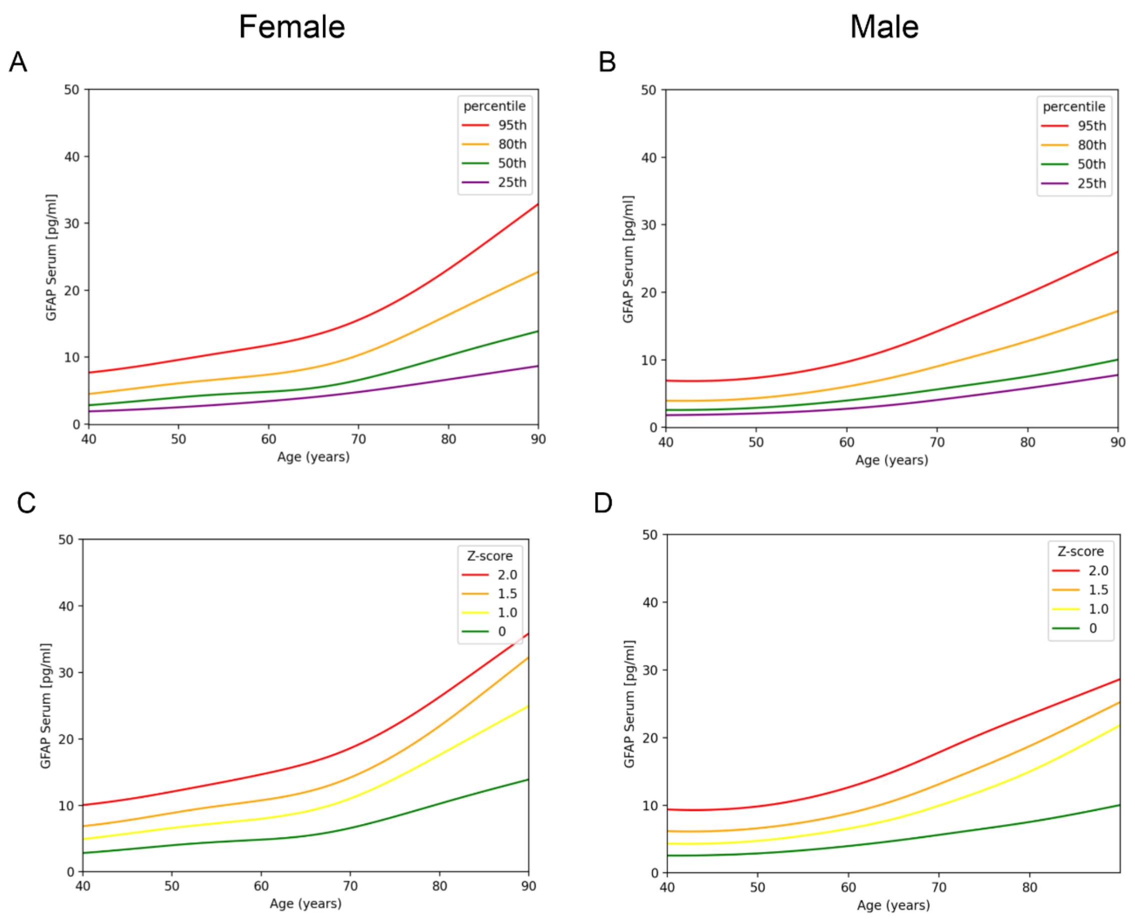


**Figure S4: Female and male serum GFAP age-dependent control reference curves from age 40 onwards**

Figure S4 **A** and **B** illustrate the serum GFAP percentiles dependent on age in females and males, respectively. In **C** and **D** the z-scores from z=0 until z=2 are according to sex and age are shown. For the modelling additive quantile regression was used. Abbreviations: GFAP; glial fibrillary acidic protein

Table S3: Age-specific GFAP z-scores according to sex in the control cohort

| Age | Female z=0 [pg/ml] | Female z=2 [pg/ml] | Male z=0 [pg/ml] | Male z=2 [pg/ml] |
|---|---|---|---|---|
| 40 | 2.8 | 10.0 | 2.57 | 9.40 |
| 45 | 3.39 | 10.9 | 2.63 | 9.40 |
| 50 | 4.00 | 12.1 | 2.89 | 9.83 |
| 55 | 4.48 | 13.3 | 3.35 | 10.9 |
| 60 | 4.84 | 14.7 | 3.97 | 12.7 |
| 65 | 5.41 | 16.3 | 4.73 | 15.0 |
| 70 | 6.59 | 18.6 | 5.62 | 17.8 |
| 75 | 8.32 | 22.0 | 6.55 | 20.7 |
| 80 | 10.3 | 26.3 | 7.53 | 23.4 |
| 85 | 12.1 | 31.1 | 8.71 | 26.1 |

Abbreviations: AD, Alzheimer's disease; Con, control; GFAP, Glial fibrillary acidic protein

**Influence of age, sex and BMI on GFAP levels in a linear model**

To test the influence of age, sex and BMI (which was available from 560 patients) on serum GFAP levels in the control group we performed a multiple linear regression analysis with GFAP serum levels as outcome and age, sex and BMI as covariates. The model had an overall goodness of fit of R2 = 0.27. All variables, age (<0.0001), sex (p=0.0002) and BMI (p=0.0008) displayed a significant influence. However, only age showed a very strong contribution with a remaining R2 of 0.0045 when age as a variable was removed. Both sex and BMI displayed very weak contributions (remaining R2: 0.25 and 0.26, respectively). For this reason we integrated only age in the main model. Due to a non-significant trend in older individuals for higher levels in females we also performed a separate analysis for females and males (see S2-S4 and table S3).

Table S4: ROC analysis Control vs. AD; Cut-off, sensitivity and specificity of the different stratified age groups

| Age group | Cut-off in [pg/ml] | Sensitivity (95% CI) in % | Specificity (95% CI) in % |
|---|---|---|---|
| 51-60 | 7.4 | 95 (84-99) | 91 (86-94) |
| 61-70 | 8.4 | 89 (80-94) | 86 (81-90) |
| 71-80 | 9.0 | 89 (83-93) | 67 (60-73) |
| 81-90 | 12.3 | 79 (65-89) | 62 (48-75) |

Abbreviations: AD, Alzheimer's disease; GFAP, Glial fibrillary acidic protein

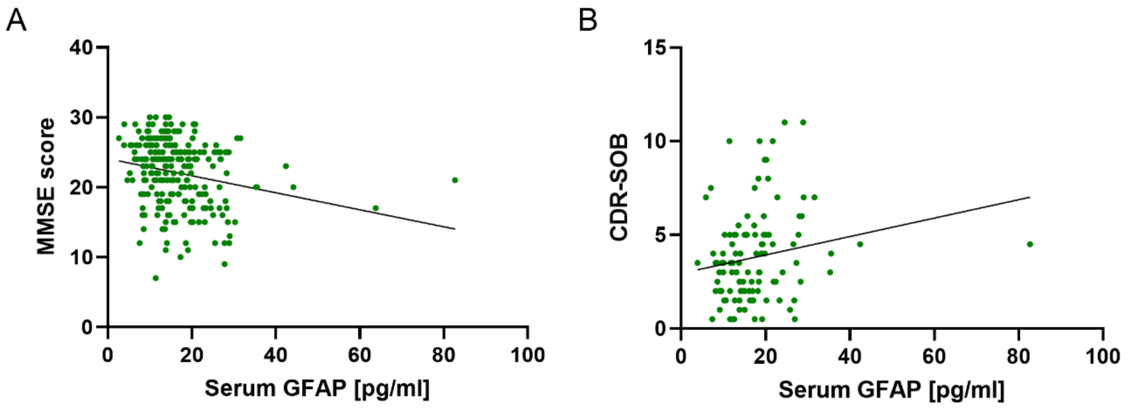


**Figure S5: Correlation of serum GFAP with cognitive scores**

(**A**) displays the correlation of serum GFAP with the MMSE score depicting a correlation r of (-0.27 (-0.39--0.14), p<0.0001). Figure S1 **B** shows the serum GFAP correlation with the CDR-SOB (CDR-SOB (r=0.22 (0.03-0.40), p=0.01). Abbreviations: CDR-SOB; Clinical dementia rating sum of boxes; GFAP, glial fibrillary acidic protein; MMSE, Mini Mental State Examination

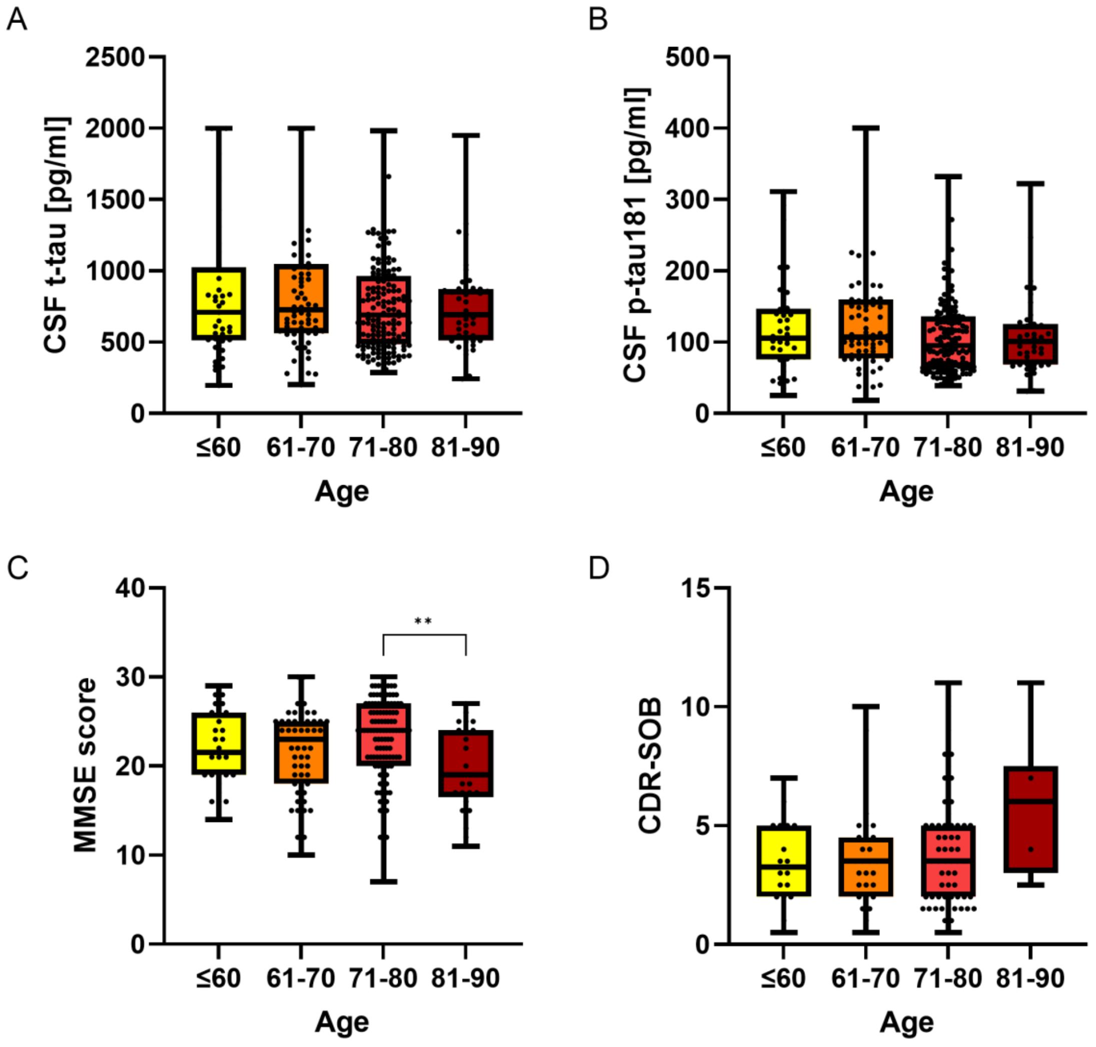


**Figure S6: CSF t-tau, p-tau181 and cognitive scores in the different age groups**

Figure S2 **A** and **B** illustrate the distribution of CSF t-tau and p-tau181 levels in the different age groups. No significant difference was detected. **C** and **D** display the cognitive score results by age group with the MMSE in C and the CDR-SOB in D. Here the MMSE score showed significantly lower scores in the older age group. Using the CDR-SOB a trend to higher levels in the oldest age group is visible. Abbreviations: CDR-SOB; Clinical dementia rating sum of boxes; MMSE, Mini Mental State Examination; p-tau, phosphorylated t-tau, total-tau.

**AD patient stratification**

To stratify the AD group into AD-dementia (ADD) and AD with MCI (AD-MCI), the clinical dementia rating (CDR) more specifically the CDR-SOB was used [1, 2]. Patients with a CDR SOB below 2.5 were considered AD-MCI (Figure S2A) [3]. In a second more crude approach the MMSE was used for the stratification. Patients below the cut off of 25 were considered ADD and AD-MCI with a score of 25 or 26 (Figure S2B) [4].

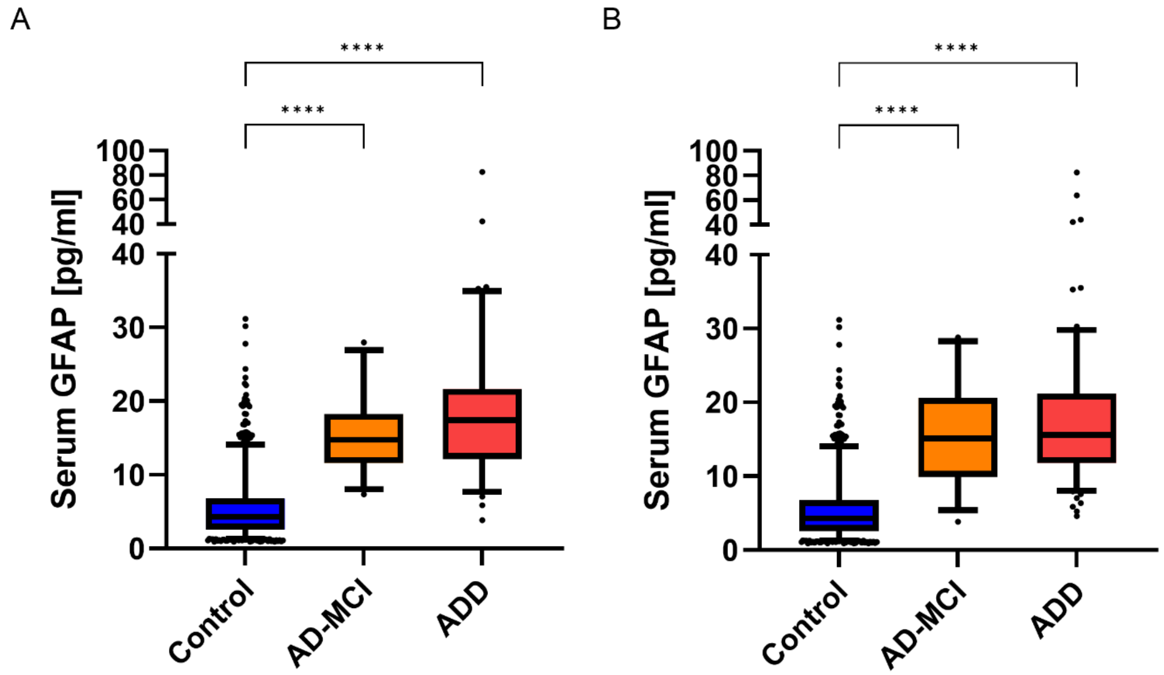


**Figure S7: Serum GFAP levels according to cognitive impairment stage**

In S3 **A** the AD group was stratified according to the results of the CDR test (AD-MCI: CDR-SOB values <2.5 and ADD ≥2.5). In **B** the AD patients were stratified according to the MMSE (AD-MCI: MMSE 25-26 and ADD≤24). Both stratifications show significantly elevated serum GFAP levels in the AD-MCI and ADD group compared to the control patients. No significant difference was found between the AD-MCI and ADD groups. ****, p<0.0001. Abbreviations: ADD, Alzheimer's disease dementia; AD-MCI, Alzheimer's disease mild cognitive impairment; CDR-SOB; Clinical dementia rating sum of boxes; GFAP, glial fibrillary acidic protein; MMSE, Mini Mental State Examination

**Members of the ALS registry study group:**

Dr. med. Alber, B., Klinikum Günzburg, Department of Neurology

Prof. Dr. med. Arnold G., Klinikum Sindelfingen-Boeblingen, Department of Neurology

Dr. med. Baier H., ZFP Suedwuerttemberg, Department of Epileptology

Prof. Dr. med. Baezner H., Katharinenhospital Stuttgart, Department of Neurology

Dr. med. Beattie J., Ostalb-Klinikum Aalen, Department of Neurology

Dr. med. Behne F., ZFP Suedwuerttemberg, Department of Epileptology

Prof. Dr. med. Bengel D., Oberschwabenklinik Ravensburg, Department of Neurology

Dr. med. Boertlein A.,Katharinenhospital Stuttgart, Department of Neurology

Dr. med. Dempewolf, S., Department of Neurology, Ludwigsburg

Prof. Dr. med. Dettmers C., Schmieder Kliniken Konstanz

Prof. Dr. med. Freund, W., Praxis Biberach

Dr. med. Gold H.-J., Klinikum am Gesundbrunnen Heilbronn, Department of Neurology

Prof. Dr. med. Wick, W., University of Heidelberg, Department of Neurology

Prof. Dr. med. Hecht M., Bezirkskrankenhaus Kaufbeuren, Department of Neurology

Dr. med. Heimbach B., University of Freiburg, Department of Neurology

Prof. Dr. med. Herting B., Diakonie-Klinikum Schwaebisch Hall, Department of Neurology

Prof. Dr. med. Huber R., Klinikum Friedrichshafen, Department of Neurology

Prof. Dr. med. Huelser P.-J., Fachklinik Wangen, Department of Neurology

Dr. med. Engelberger, M., Kliniken Landkreis Heidenheim, Department of Neurology

Dr. med. Kaspar A., Oberschwabenklinik Ravensburg, Department of Neurology

Prof. Dr. med. Kimmig H., Kliniken Schwenningen, Department of Neurology

Prof. Dr. med. Zeller D., University of Würzburg, Department of Neurology

Prof. Dr. med. Kloetzsch C., Hegau-Bodensee-Klinikum Singen, Department of Neurology

Prof. Dr. med. Klopstock, T., LMU München, Department of Neurology

Dr. med. Kohler, A., Klinikum am Gesundbrunnen Heilbronn, Department of Neurology

PD Dr. med. Lichy C., Klinikum Memmingen, Department of Neurology

Prof. Dr. med. Lindner A., Marienhospital Stuttgart, Department of Neurology

PD Dr. med. Buttmann M., Caritas Krankenhaus, Bad Mergentheim, Department of Neurology

Dr. med. Meyer A., Weissenau, Department of Neurology

Prof. Dr. med. Synofzik, M., Universitaetsklinikum Tuebingen, Department of Neurology

Dr. med. Clauer-Bredt, M., Christophsbad Goeppingen, Department of Neurology

Prof. Dr. med. Naumann M., Klinikum Augsburg, Department of Neurology and Neurophysiology

Dr. med. Demuth, K., Vinzenz von Paul Hospital, Rottweil, Department of Neurology

PD Dr. med. Neuhaus O., Kliniken Landkreis Sigmaringen, Department of Neurology

Prof. Dr. med. Neusch C., Praxis EMSA Singen

Prof. Dr. med. Niehaus L., Department of Neurology, Winnenden

PD Dr. med. Jüttler E., Ostalb-Klinikum Aalen, Department of Neurology

Dr. med. Ratzka P., Klinikum Augsburg, Department of Neurology and Neurophysiology

Prof. Dr. med. Förch, C., Department of Neurology, Ludwigsburg

Prof. Dr. med. Gasser, T., Universitaetsklinikum Tuebingen, Department of Neurology

Dr. med. Schweigert B., Caritas Krankenhaus, Bad Mergentheim, Department of Neurology

PD Dr. K. Althaus, Christophsbad Goeppingen, Department of Neurology

Prof. Dr. med. Reinhard M., Kliniken Esslingen, Department of Neurology

Dr. med. Trottenberg T., Department of Neurology, Winnenden

Oberstarzt Dr. med. Metrikat J., Bundeswehrkrankenhaus Ulm, Department of Neurology

Prof. Dr. med. Weiler M., University of Heidelberg, Department of Neuology

Prof. Dr. med. Volkmann J., University of Wuerzburg, Department of Neurology

Prof. Dr. med. Hamann G., Klinikum Guenzburg, Department of Neurology

Prof. Dr. med. Pinkhardt, E., Klinikum Kempten, Department of Neurology

Dr. med. Andres, Kreiskliniken Reutlingen, Department of Neurology

Prof. Dr. med. Dorothee Lulé, Ulm University, Department of Neurology

Prof. Dr. med. Opherk C., Klinikum am Gesundbrunnen Heilbronn, Department of Neurology

Prof. Dr. med. Hemmer B., TU München, Department of Neurology

Prof. Dr. med. Weiller C., University of Freiburg, Department of Neurology

Prof. Dr. med. Lingor P., TU München, Department of Neurology

Dr. med. Schläger, A., Kliniken Esslingen, Department of Neurology

Prof. Dr. med. Jöbges, M., Schmieder Kliniken Konstanz

Prof. Dr. med. Nenad Vasic, Allgemeinpsych., Christophsbad Göppingen

Dr. Felix Geser, Christophsbad Göppingen

Dr.med. Karsten Henkel, Gerontopsychiatrie Christophsbad Göppingen

Dr. med. Martin Zinkler, Kliniken Heidenheim

Dr.med. Ralf Kozian, Gerontopsych.Vinzenz von Paul Hospital, Rottweil

Prof. Dr. med. Dr. phil. Martin Bürgy, Allgemeinpsych Zentrum f. Seelische Gesundheit, Klinikum Stuttgart, Bad Cannstatt

Priv.-Doz. Dr. med. Christine Thomas, Gerontopsych Zentrum f. Seelische Gesundheit,Klinikum Stuttgart, Bad Cannstatt

Dr.med. Stefan Spannhorst, Zentrum für Seelische Gesundheit,Klinikum Stuttgart, Bad Cannstatt

Priv.-Doz. Dr. med. Matthias Munk Unilklinik Tübingen

Prof.Dr. Christoph Laske Uniklinik Tübingen

PD Dr.med.Daniel Schüpbach ZfP Weinsberg, Klinikum am Weissenhof, Allgemeinpsych West

PD Dr.med.Heinz Grunze ZfP Weinsberg, Klinikum am Weissenhof, Allgemeinpsych Ost

Dr. Rainer Schaub, Gerontopsych ZfP Weinsberg, Klinikum am Weissenhof

Dr. Jochen Gebhardt, Gerontopsychiatrie ZfP Wiesloch

Dr.med. Hubertus Friederich, Gerontopsych. ZFP Zwiefalten

Dr. Matthias Köhler, Gerontopsych. ZFP Zwiefalten

Dr.med.Alex Gogolkiewicz, Allgemeinpsych.ZfP Zwiefalten

Prof.Dr. med. Andreas Joos Kliniken Schmieder Gailingen

Prof. Dr. med. Max Schmauß BKH Augsburg

Dr.med.Jessica Baumgärtner BKH Augsburg

Prof. Riepe, Gerontopsych.BKH Günzburg , Geontopsych

Prof. Becker, Allgemeinpsych.BKH Günzburg,  Psych.II

Dr. Andreas Küthmann Bezirkskrankenhaus Memmingen

Dr. Raimund Steber Bezirkskrankenhaus Memmingen

Dr. Ivona Gecui Bezirkskrankenhaus Memmingen

Univ. Prof. Dr. med. Elmar Etzersdorfer Furtbachkrankenhaus, Stuttgart

Dr.med Alexandros Michaelides, Furtbachkrankenhaus, Stuttgart

Prof. Dr. med. Bernhard Connemann, Uniklinik Ulm, Psych. III

Andreas Raether, Gerontopsychiatrie, ZfP Winnenden
